# Sex-Specific TMPRSS2 Response and Reduced Peripheral RNA Concentration Following AstraZeneca COVID-19 Vaccination in Nigeria

**DOI:** 10.64898/2026.06.21.26356163

**Authors:** Erens Spiff Ekprikpo, Stella Urekweru Ken-Ezihuo, Beauty Eruchi Echonwere-Uwikor, Zaccheaus Awortu Jeremiah

## Abstract

**Background:** ChAdOx1 nCoV-19 remains a cornerstone COVID-19 vaccine in sub-Saharan Africa, yet population-specific molecular responses are understudied. We examined peripheral blood ACE2 and TMPRSS2 expression, total RNA concentration, and coagulation indices in Nigerians ≥6 months post-vaccination.

**Methods:** In a case-control study in Port Harcourt, Nigeria, 51 ChAdOx1-vaccinated adults and 51 age/sex-matched unvaccinated controls provided venous blood for RNA extraction, qRT-PCR, and coagulation assays. Multivariable linear models assessed effects of vaccination, sex, and age on molecular parameters.

**Results:** Vaccinated participants had 37% lower total RNA concentration than controls (4.02 ± 0.09 vs 6.38 ± 0.14 ng/μL, p<0.0001). ACE2 and TMPRSS2 expression did not differ by vaccination status overall. However, TMPRSS2 showed a significant sex-by-treatment interaction (p=0.011): vaccinated females had higher expression than vaccinated males. GAPDH expression varied by vaccination status and showed a three-way interaction with sex and age (p=0.027). Coagulation indices were unchanged.

**Conclusions:** At ≥6 months post-ChAdOx1, Nigerians show reduced peripheral blood RNA without sustained ACE2/TMPRSS2 upregulation. The sex-specific TMPRSS2 pattern suggests hormone×vaccine interactions previously unreported in African cohorts and highlights the need for sex-disaggregated molecular surveillance. Region-specific reference gene validation is recommended for Nigerian transcriptomic studies.

## Introduction

COVID-19 vaccines have been deployed at unprecedented speed, yet molecular data from tropical, sub-Saharan African populations remain sparse. ChAdOx1 nCoV-19 (AstraZeneca) is widely used in Nigeria and across Africa, but most post-vaccination gene expression studies derive from cell lines or European cohorts [1, 2]. SARS-CoV-2 cell entry requires angiotensin-converting enzyme 2 (ACE2) and transmembrane serine protease 2 (TMPRSS2) [3, 4]. Early studies suggested vaccines might transiently alter these entry factors, with potential implications for host susceptibility [5].

ACE2 is expressed in lung, heart, kidney, and gastrointestinal tissues and functions within the renin-angiotensin system [6]. TMPRSS2 primes viral spike proteins and is androgen-regulated, a mechanism linked to male-biased COVID-19 severity [7, 8]. Recent reports describe transient ACE2 upregulation and TMPRSS2 downregulation after vaccination, but long-term patterns in African populations are unknown [5].

Given Nigeria’s high ChAdOx1 uptake and distinct genetic and environmental context, population-specific molecular surveillance is needed. We assessed peripheral blood ACE2 and TMPRSS2 expression, total RNA concentration, and coagulation profiles in Nigerian adults ≥6 months after ChAdOx1 vaccination, testing for effects of sex and age.

## Materials and Methods

### Study design and participants

This case-control study was conducted in Port Harcourt, Nigeria, from March to June 2024. We enrolled 51 adults who had received ≥1 dose of ChAdOx1 nCoV-19 ≥6 months prior and 51 age-and sex-matched unvaccinated controls aged 18–65 years. Exclusion criteria included pregnancy, active infection, autoimmune disease, anticoagulant therapy, or known hemostatic disorders. Sample size was calculated using G*Power (version 3.1.9.4) for a medium effect size (Cohen’s d = 0.5), α = 0.05, power = 0.80.

### Sample Collection and Processing

Venous blood was collected in EDTA tubes and stored at −80°C. Total RNA was extracted from whole blood using the Zymo Quick-RNA Plus Isolation Kit (Zymo Research, USA). RNA concentration and purity (A260/A280 ratio) were quantified by NanoDrop.

### qRT-PCR

cDNA was synthesized using the Bio-Rad iScript cDNA Synthesis Kit. qRT-PCR for ACE2, TMPRSS2, and GAPDH was performed with Radiant SYBR Green 1-Step Lo-Rox qPCR Kit (Alkali Scientific, USA) on a Bio-Rad CFX96 Real-Time PCR Detection System. All reactions included no-template controls and were run in duplicate. Cycle threshold (CT) values were recorded; higher CT indicates lower gene expression. GAPDH was assessed as a candidate reference gene. Primer sequences are available upon request.

### Coagulation Assays

Prothrombin time (PT), activated partial thromboplastin time (APTT), and international normalized ratio (INR) were measured using standard hematology protocols on an automated coagulometer.

### Statistical Analysis

Independent t-tests compared vaccinated and unvaccinated groups. Multivariable linear regression tested effects of vaccination status, sex, age, and their interactions on log-transformed outcomes. Two-way and three-way ANOVA examined interaction effects. Analyses were performed using SAS v9.4 and JMP v14.3. Significance was set at p<0.05.

### Ethics Statement

The study was approved by the Rivers State Hospital Management Board Research Ethics Committee (approval number: RSHMB/RSHREC/2024/113). All procedures were performed in accordance with the ethical standards of the institutional research committee and the Declaration of Helsinki. Participants were recruited from March 2024 to June 2024. They were provided with a detailed explanation of the study purpose, procedures, potential risks, and benefits. Informed consent was obtained from all participants after this explanation. The consent process was conducted in accordance with the approved protocol.

### Data Availability

All relevant data are within the manuscript and its Supporting Information files. Raw qPCR CT values and de-identified participant data are available from the corresponding author upon reasonable request.

## Results

Demographic characteristics were comparable between groups (Table 1). Mean age was 38.8 ± 6.3 years for vaccinated and 34.8 ± 10.3 years for unvaccinated participants.

**Table 1.** Demographic characteristics of study participants (n=102).

|  | Total | Vaccinated (Test) | Unvaccinated (Control) |
| --- | --- | --- | --- |
| Characteristic | N (%) | n (%) | n (%) |
| **Sex** |  |  |  |
| Female | 45 (44.1) | 18 (17.7) | 27 (26.5) |
| Male | 57 (55.9) | 33 (32.4) | 24 (23.5) |
| **Age Group (years)** | 29 (28.4) | 5 (4.9) | 24 (23.5) |
| <30 | 51 (50.0) | 35 (34.3) | 16 (15.7) |
| 30-44 | 22 (21.6) | 11 (10.8) | 11 (10.8) |
| 45+ |  |  |  |
| **Mean ± SD** | 36.8 ± 8.7 | 38.8 ± 6.28 | 34.8 ± 10.3 |

Vaccinated participants had significantly lower peripheral blood total RNA concentration than controls (4.02 ± 0.09 vs 6.38 ± 0.14 ng/μL, p<0.0001; Table 2). ACE2 and TMPRSS2 expression did not differ significantly by vaccination status (p=0.056 and p=0.342). GAPDH expression was significantly higher in vaccinated participants (p=0.027).

**Table 2.**
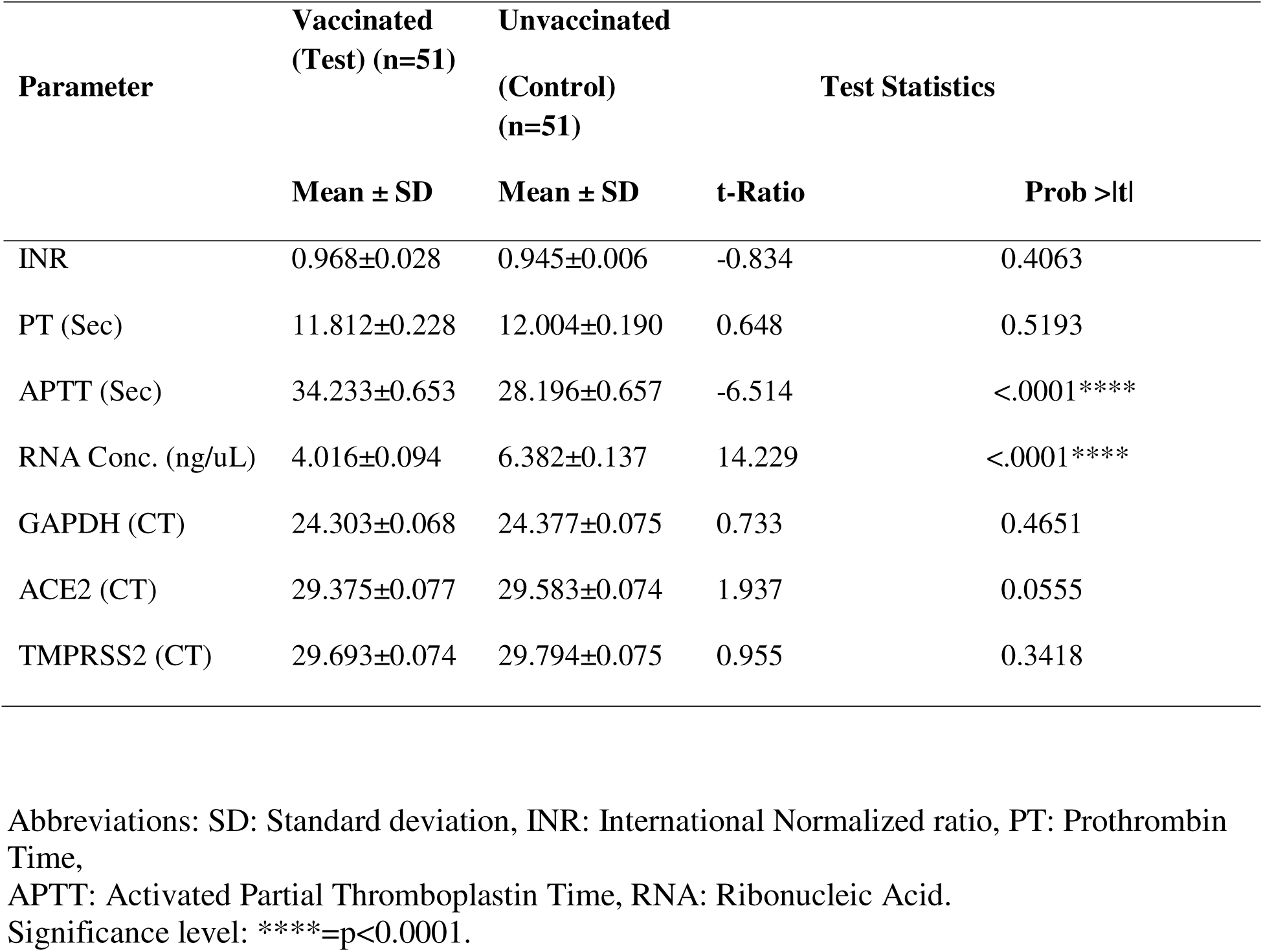
Coagulation and Molecular Parameters in ChAdOx1 nCoV-19 Vaccinated Versus Unvaccinated Subjects.

| Parameter | Vaccinated<br>(Test) (n=51) | Unvaccinated<br>(Control)<br>(n=51) | Test Statistics |  |
| --- | --- | --- | --- | --- |
| | Mean ± SD | Mean ± SD | t-Ratio | Prob >\|t\| |
| INR | 0.968±0.028 | 0.945±0.006 | -0.834 | 0.4063 |
| PT (Sec) | 11.812±0.228 | 12.004±0.190 | 0.648 | 0.5193 |
| APTT (Sec) | 34.233±0.653 | 28.196±0.657 | -6.514 | <.0001**** |
| RNA Conc. (ng/uL) | 4.016±0.094 | 6.382±0.137 | 14.229 | <.0001**** |
| GAPDH (CT) | 24.303±0.068 | 24.377±0.075 | 0.733 | 0.4651 |
| ACE2 (CT) | 29.375±0.077 | 29.583±0.074 | 1.937 | 0.0555 |
| TMPRSS2 (CT) | 29.693±0.074 | 29.794±0.075 | 0.955 | 0.3418 |

Abbreviations: SD: Standard deviation, INR: International Normalized ratio, PT: Prothrombin Time, APTT: Activated Partial Thromboplastin Time, RNA: Ribonucleic Acid. Significance level: \*\*\*\*=p<0.0001.

Multivariable models showed no significant treatment-by-age interaction (Table 4). However, a significant sex-by-treatment interaction was identified for TMPRSS2 (F=6.80, p=0.011; Table 3): vaccinated females had lower CT values, indicating higher expression, than vaccinated males. No interaction was observed for ACE2. A three-way interaction of treatment, sex, and age was significant for GAPDH (F=3.78, p=0.027; Table 5), with vaccinated females aged 30–44 showing the highest expression. These patterns are illustrated in Fig 1.

**Fig 1.**
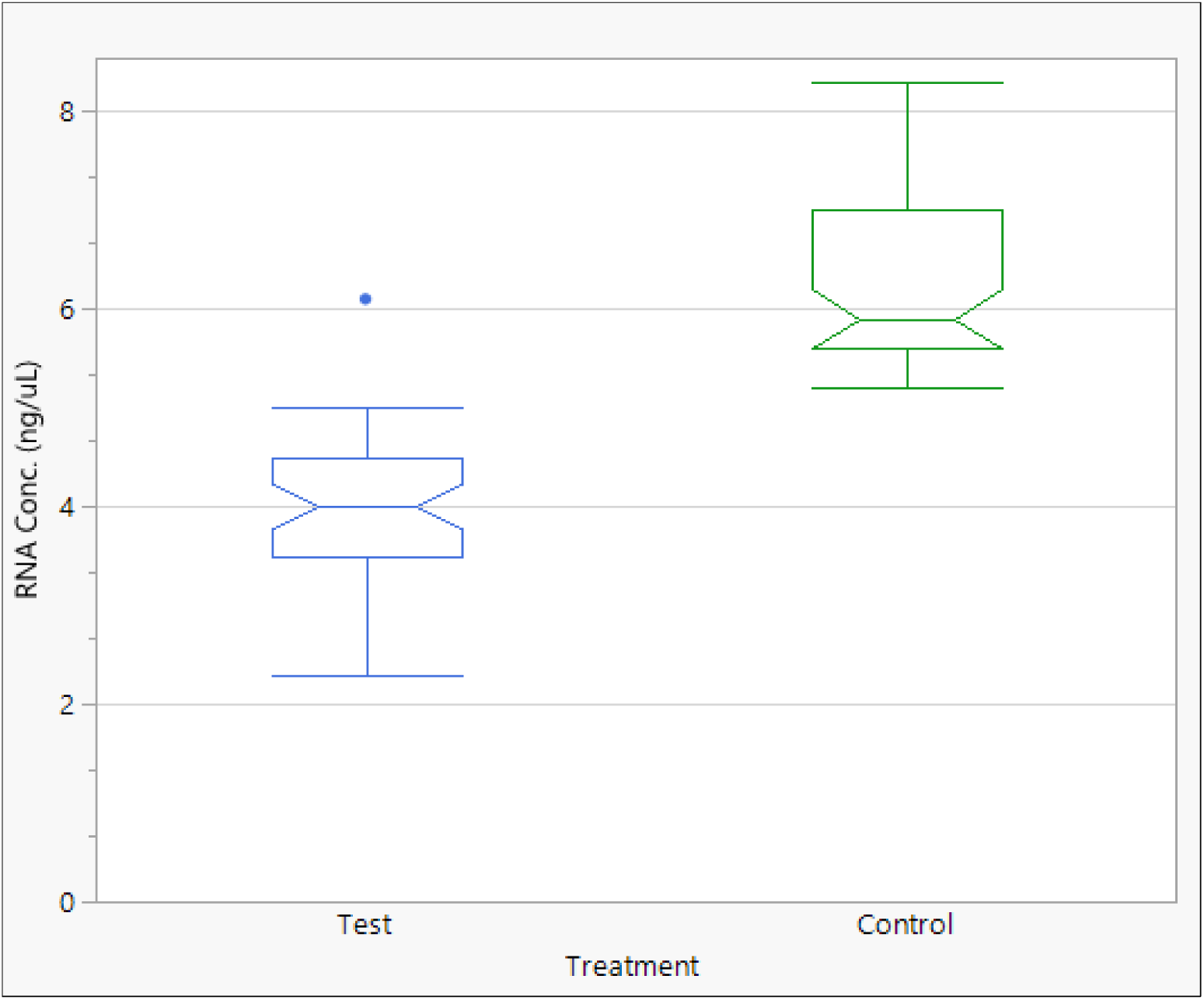
Box plots of cycle threshold (CT) values for ACE2, TMPRSS2, and GAPDH in vaccinated and unvaccinated participants, stratified by sex. Higher CT indicates lower gene expression. Horizontal line = median, box = interquartile range, whiskers = 1.5×IQR. A significant sex-by-treatment interaction was observed for TMPRSS2 (p=0.011), with vaccinated females showing lower CT (higher expression) than vaccinated males.

**Table 3.** Interaction effects of treatment and sex on molecular parameters. Mean ± SD. Values within a parameter with different superscripts differ significantly at p<0.05.

| Treatment | Sex | n | RNA Conc.<br>(ng/uL) | GAPDH<br>(CT) | ACE2<br>(CT) | TMPRSS2<br>(CT) |
| --- | --- | --- | --- | --- | --- | --- |
| | | | Mean ± SD | Mean ±SD | Mean ±SD | Mean ± SD |
| Vaccinated (Test) | Female | 18 | 4.03±0.26 | 24.36±0.15 | 29.43±0.17 | 29.39±0.15<sup>a</sup> |
| | Male | 33 | 3.80±0.20 | 24.44±0.12 | 29.42±0.13 | 29.98±0.12<sup>b</sup> |
| Unvaccinated<br>(Control) | Female | 27 | 6.39±0.16 | 24.34±0.10 | 29.64±0.11 | 29.84±0.09<sup>b</sup> |
| | Male | 24 | 6.22±0.21 | 24.46±1.12 | 29.58±0.14 | 29.79±0.12<sup>b</sup> |
| Test Statistics |  |  |  |  |  |  |
| *F-Ratio* |  |  | 0.0152 | 0.0267 | 0.0246 | 6.7961 |
| *P-value* |  |  | 0.9021<sup>ns</sup> | 0.8707<sup>ns</sup> | 0.8757<sup>ns</sup> | 0.0107** |

Abbreviations: SD: Standard deviation.
Mean ± SD within a given parameter with different superscripts are significantly different at $p < 0.05$ . Significance level: \*\*= $p < 0.01$ , ns = not significant ( $p > 0.05$ ).

**Table 4.** Interaction Effects of Treatment and Age Group on Molecular Parameters of COVID-19 Vaccinated and Unvaccinated Subjects.

| Treatment | Age Group<br>(years) | N | RNA Conc.<br>(ng/uL)<br>Mean ± SD | GAPDH<br>(CT)<br>Mean ± SD | ACE2 (CT)<br>Mean ± SD | TMPRSS2 (CT)<br>Mean ± SD |
| --- | --- | --- | --- | --- | --- | --- |
| Vaccinated<br>(Test) | <30 | 5 | 3.66±0.39 | 24.62±0.23 | 29.51±0.26 | 29.75±0.22 |
|  | 30-44 | 35 | 4.16±0.15 | 24.21±0.09 | 29.35±0.10 | 29.51±0.09 |
|  | 45<sup>+</sup> | 11 | 3.93±0.27 | 24.37±0.16 | 29.40±0.18 | 29.79±0.15 |
| Unvaccinated<br>(Control) | <30 | 24 | 6.41±0.18 | 24.40±0.10 | 29.53±0.12 | 29.79±0.10 |
|  | 30-44 | 16 | 6.39±0.21 | 24.44±0.13 | 29.56±0.14 | 29.76±0.12 |
|  | 45<sup>+</sup> | 11 | 6.11±0.29 | 24.36±0.17 | 29.74±0.19 | 29.89±0.17 |
| Test Statistics |  |  |  |  |  |  |
| *F-Ratio* |  |  | 0.6378 | 1.2943 | 0.3727 | 0.3393 |
| *P-Value* |  |  | 0.5308<sup>ns</sup> | 0.2791<sup>ns</sup> | 0.6899<sup>ns</sup> | 0.7132<sup>ns</sup> |

Abbreviations: SD: Standard deviation. Significance level: ns= Not significant (p>0.05).

**Table 5.** Interaction effects of treatment, sex, and age group on molecular parameters. Mean ± SD. Values within a parameter with different superscripts differ significantly at p<0.05.

| Treatment | Sex | Age Group | n | RNA Conc. (ng/uL) | GAPDH (CT) | ACE2 (CT) | TMPRSS2 (CT) |
| --- | --- | --- | --- | --- | --- | --- | --- |
|  |  | (years) |  | Mean ± SD | Mean ± SD | Mean ± SD | Mean ± SD |
| Vaccinated<br>(Test) | Female | <30 | 2 | 3.95±0.60 | 24.42±0.36<sup>abc</sup> | 29.53±0.40 | 29.60±0.35 |
|  |  | 30-44 | 12 | 4.42±0.25 | 24.08±0.15<sup>b</sup> | 29.37±0.16 | 29.17±0.14 |
|  |  | 45<sup>+</sup> | 4 | 3.73±0.43 | 24.59±0.25<sup>abc</sup> | 29.39±0.28 | 29.40±0.25 |
|  | Male | <30 | 3 | 3.37±0.49 | 24.83±0.29<sup>a</sup> | 29.50±0.32 | 29.90±0.28 |
|  |  | 30-44 | 23 | 3.91±0.18 | 24.34±0.11<sup>abc</sup> | 29.34±0.12 | 29.85±0.10 |
|  |  | 45<sup>+</sup> | 7 | 4.13±0.32 | 24.15±0.19<sup>abc</sup> | 29.41±0.21 | 30.18±0.19 |
| Unvaccinated<br>(Control) | Female | <30 | 10 | 6.32±0.27 | 24.42±0.16<sup>abc</sup> | 29.51±0.18 | 29.84±0.16 |
|  |  | 30-44 | 9 | 6.37±0.28 | 24.53±0.17<sup>a,c</sup> | 29.76±0.19 | 29.92±0.16 |
|  |  | 45<sup>+</sup> | 8 | 6.49±0.30 | 24.06±0.18<sup>bc</sup> | 29.64±0.20 | 29.76±0.17 |
|  | Male | <30 | 14 | 6.50±0.23 | 24.38±0.14<sup>abc</sup> | 29.54±0.15 | 29.75±0.13 |
|  |  | 30-44 | 7 | 6.41±0.32 | 24.35±0.19<sup>abc</sup> | 29.36±0.21 | 29.60±0.19 |
|  |  | 45<sup>+</sup> | 3 | 5.73±0.49 | 24.65±0.29<sup>abc</sup> | 29.85±0.32 | 30.01±0.28 |
| Test Statistics |  |  |  |  |  |  |  |
| F-Ratio |  |  |  | 1.9382 | 3.7792 | 0.4862 | 0.7194 |
| P-Value |  |  |  | 0.1499 | 0.0265* | 0.6165 | 0.4898 |

Abbreviations: SD: Standard deviation.
Mean ± SD within a given parameter with different superscripts are significantly different at $p < 0.05$ . Significance level: ns= Not significant ( $p > 0.05$ ).

Coagulation indices PT and INR did not differ between groups. APTT was significantly prolonged in vaccinated participants (34.23 ± 0.65 vs 28.20 ± 0.66 sec, p<0.0001), but remained within the clinical reference range (Table 2).

## Discussion

Six months after ChAdOx1 nCoV-19 vaccination, Nigerian adults showed a 37% reduction in peripheral blood total RNA concentration without sustained upregulation of ACE2 or TMPRSS2. This contrasts with early cell-line studies reporting transient ACE2 increases post-vaccination and suggests that initial transcriptional responses do not persist at later timepoints in whole blood in this population [5].

The reduced RNA concentration likely reflects immune contraction following the post-vaccination expansion phase. Activated leukocytes are a major source of RNA in whole blood, and their decline during immune homeostasis could explain the observed decrease. We did not perform differential blood counts, so this mechanism remains to be tested directly. Importantly, there is no evidence that ChAdOx1 destabilizes RNA.

The most notable finding was the sex-specific TMPRSS2 response. Vaccinated females exhibited higher TMPRSS2 expression than vaccinated males. TMPRSS2 is androgen-regulated and has been implicated in the male bias of severe COVID-19 [7,8]. Our data raise the possibility that ChAdOx1 interacts with sex hormone pathways in Africans, a pattern not previously documented. Sex differences in vaccine responses are well established for influenza and HPV vaccines, where females often mount stronger responses [9]. Whether the TMPRSS2 signal has clinical relevance for SARS-CoV-2 susceptibility or future coronavirus vaccines requires dedicated studies incorporating hormone measurements and immune phenotyping.

The observed variability in GAPDH, a common reference gene, and its interaction with sex and age, provides a methodological caution. Housekeeping genes are not uniformly stable across populations or physiological states. Transcriptomic studies in Nigerian cohorts should validate reference genes locally rather than assume GAPDH stability.

## Limitations

Limitations include the single, late timepoint (>6 months), which precludes analysis of acute-phase responses. Whole blood captures mixed cell populations; cell-type-specific changes cannot be resolved here. We did not measure antibody titers, breakthrough infections, sex hormone levels, or perform differential blood counts. The sample size is powered to detect medium-to-large effects.

## Implications

This study provides baseline molecular surveillance for ChAdOx1 in a tropical African setting. The absence of sustained ACE2/TMPRSS2 upregulation is consistent with clinical safety monitoring in Nigeria. The sex-specific TMPRSS2 finding generates a testable hypothesis regarding hormone×vaccine interactions in African populations. Equity in vaccine science requires measuring these differences rather than extrapolating from other regions.

## Conclusion

At ≥6 months post-ChAdOx1 nCoV-19 vaccination, Nigerian adults exhibited a 37% reduction in peripheral blood total RNA concentration compared to unvaccinated controls, with no significant alteration in ACE2 or TMPRSS2 expression overall. A significant sex-by-treatment interaction for TMPRSS2 indicates higher expression in vaccinated females than vaccinated males. Coagulation indices remained within clinical reference ranges. These findings underscore the need for sex-disaggregated molecular analyses in African vaccine studies. Future research should examine acute-phase dynamics, immune cell subsets, and clinical correlates of RNA reduction. Validation of reference genes for Nigerian transcriptomic work is recommended.

## Acknowledgments

We thank the participants and staff at Rivers State University Teaching Hospital. No specific funding was received for this study.

